# Patient acceptance of preventive antibiotic treatment for tuberculosis: a qualitative study

**DOI:** 10.64898/2026.08.07.26359686

**Authors:** Mélanie Antunes, Rubin Rose-Key, Ellen Steward, Sammy Assayad, Mahdad Noursadeghi, Luis Loria Rebolledo, Elise Crayton, Rishi K Gupta

**Author notes:** Correspondence: Dr Rishi K Gupta, UCL Respiratory, 1st floor Rayne building, 5 University Street, WC1E 6JF; r. Contributed equally (Elise Crayton, Rishi K Gupta).

## Abstract

**Background:** Tuberculosis (TB) preventive treatment is a key component of TB control in low-incidence settings, but uptake remains low.

**Objective:** To explore factors influencing patient decisions to accept or decline preventive treatment for TB infection.

**Design:** Qualitative study using semi-structured interviews, analysed with the Theoretical Domains Framework (TDF).

**Setting:** Routine clinical TB prevention services at two hospitals in London, UK.

**Participants:** Adults (≥18 years) diagnosed with TB infection and offered preventive treatment.

**Methods:** Semi-structured interviews were audio-recorded and transcribed verbatim. The TDF was applied to analyse transcripts using a combined inductive thematic analysis and deductive framework approach. Themes were also mapped to the Capability, Opportunity and Motivation model of Behaviour (COM- B) domains.

**Results:** Twenty-five participants (median age 34 years; 64% male) were included; 56% accepted treatment, 28% declined, and 16% were undecided. Influences on decision-making mapped to 12 of 14 TDF domains. Facilitators of treatment acceptance included perceived risk of TB (beliefs about consequences), desire to protect others’ health (social influences and goals), confidence in treatment adherence (beliefs about capabilities), and routine integration strategies (behavioural regulation). Barriers to treatment acceptance included low perceived individual risk and doubts about necessity or effectiveness (beliefs about consequences), concerns about side effects and treatment burden (environmental context and resources) and anticipated stigma (social influences). Knowledge had a mixed influence, primarily shaping perceived necessity, but did not determine decisions alone. Social, environmental and emotional factors further influenced decisions, with participants balancing anticipated benefits against perceived burden and uncertainty across multiple interacting domains.

**Conclusions:** Decisions to accept preventive TB treatment are driven by interacting capability, opportunity and motivation factors rather than knowledge alone. Interventions to improve shared decision making should address perceived risk, treatment burden, self-efficacy and social influences.

**Registration:** Clinicaltrials.gov (NCT07024836)

**Strengths and limitations of this study:**

- Use of semi-structured interviews enabled in-depth exploration of patient decision-making about tuberculosis (TB) preventive treatment in routine clinical services.
- The Theoretical Domains Framework (TDF) provided a systematic behavioural framework for coding and interpreting influences on treatment acceptance, with subsequent mapping to Capability, Opportunity and Motivation model of Behaviour (COM-B) domains.
- Recruitment included representative adults offered preventive treatment through different screening pathways and included participants who accepted, declined, or remained undecided about treatment.
- Data were collected from two London hospital sites; future studies could examine generalisability to other settings, including higher TB incidence settings, and those with different models of TB preventive care.

## Introduction

Tuberculosis (TB) remains the leading cause of death from an infectious disease globally, with an estimated 10.7 million cases and 1.2 million deaths in 2024^1^. Preventive antibiotic therapy for individuals at increased risk is a central pillar of the WHO End TB Strategy and is particularly important for countries progressing toward pre elimination^2^. In countries with low TB incidence, people at elevated risk include recent close contacts of infectious TB cases, recent migrants from high-incidence countries, and individuals undergoing screening prior to immunosuppression^3^.

TB infection is defined as a persistent immune response to *M. tuberculosis* antigens in the absence of TB disease. Tests for TB infection provide evidence of immune memory for *M. tuberculosis*, but have limited positive predictive value for future disease - typically <5% over two years in adult contacts and migrants. Prognostic ability is limited because they do not distinguish persistent infection from historical exposure and resulting immune memory^3^. Limited positive predictive value results in a high number needed to treat to prevent incident disease, undermining the efficiency of screening programmes.

Limited perceived individual-level risk may also influence patient decisions, contributing to low uptake of preventive treatment, estimated as only ∼25% in UK migrant screening data^4,5^. Previous studies exploring factors affecting acceptance have suggested that knowledge of TB and TB infection, information from healthcare workers and interpretation of test results impact uptake^6–8^. However, these studies have mostly been done in high burden countries. One qualitative study done in the UK focused on the impact of the COVID-19 pandemic on patient perceptions of TB infection^7^. Hence, the evidence base remains sparse; little is known about how people weigh risks, benefits, and uncertainties when deciding whether to start treatment. An improved understanding of how patients make decisions on whether to accept preventive therapy could enable improved pre-treatment counselling and adherence strategies, and inform diagnostic and therapeutic innovations in TB prevention.

The decision to start a treatment, and subsequent adherence and persistence with this, is a behaviour. Use of behavioural science frameworks can enhance our understanding of a behaviour and offer insight into avenues for change. Developed through expert consensus, the Theoretical Domains Framework (TDF) is one such framework, which facilitates a holistic exploration of the individual, socio-cultural and environmental factors that drive behaviour^9,10^. The TDF consists of 14 domains spanning capability, opportunity and motivational influences on behaviour and provides more granular description of the six Capability, Opportunity and Motivation model of Behaviour (COM-B) domains. Use of this framework can underpin evidence-based selection of intervention components to change behaviour.

We conducted a qualitative study using semi structured interviews among adults offered preventive treatment for TB infection in routine UK clinical services. We applied the TDF to support investigation of the factors influencing acceptance of preventative TB treatment. Our aim was to explore the factors influencing their acceptance or decline of treatment and to generate insights that may inform better communication, decision-support tools, and targets for new diagnostics and treatment regimens in TB infection.

## Methods

### Study overview

In this prospective cohort quotative study, we recruited adults (aged ≥18 years) who had tested positive for TB infection and offered preventive treatment in routine clinical services at two London hospitals. The study was conducted with the aim of holistically exploring barriers and enablers to treatment uptake, and to support development of a subsequent discrete choice experiment to quantitatively examine determinants of treatment acceptance. The study was approved by the London - Brighton & Sussex Research Ethics Committee (25/LO/0125) and was registered on clinicaltrials.gov (NCT07024836). This study is reported in line with the consolidated criteria for reporting qualitative research (COREQ)^11^.

### Recruitment

Eligible participants included migrants from countries with high TB incidence (entered UK within 5 years), people with recent contact with a person with TB disease, and people eligible for TB infection screening prior to starting immunosuppression or through occupational health testing. Consecutive clinic attendees who met the eligibility criteria were invited to take part by clinical staff. We sought to include a diverse range of participants, representative of the target population including those who accepted and those who declined treatment. All potential participants were provided with a participant information leaflet and gave informed consent prior to participation. Participants were compensated £10 for their time, paid by an online voucher. We aimed to recruit between 20 and 30 participants until we reached thematic saturation among both treatment acceptors and decliners (*i.e.*, where no new information emerged from the analysis of subsequent interviews), in line with Francis et al’s (2010) 10+3 rule^12^.

### Interviews

Interviews were conducted either in-person or online (in a private space), according to participant preference, and were digitally audio-recorded and transcribed verbatim. For non-English speakers, telephone translation services were available. Interviews were conducted by a male clinical research nurse (RRK or SA, both MSc), trained in qualitative interviewing through short-course training and through this study by a behavioural scientist (EC). No relationships existed between interviewers and interviewees before the study, and no repeat interviews were conducted. Field notes were not recorded. Transcripts were not shared with participants and they were not asked to feed back on findings. No participants dropped out after consenting.

A semi-structured interview guide (Appendix) was used to ensure essential topics were covered in each interview. The interview guide was developed with input from the research team, patient and public involvement panel, and a literature review including previous qualitative studies^6–8^. The interview guide included questions and prompts to explore beliefs about TB disease and infection, perceptions of future TB risk and preventive treatment, treatment preferences, side effects, motivations, and expected treatment outcomes, encouraging participants to reflect on the trade-offs between these factors in their decision-making.

### Analysis

Pseudonymised Interview transcripts were imported into NVivo (version 14) for analysis. As advocated by Atkins et al (2017), a combined deductive framework and inductive thematic approach was taken^10^. Analysis involved an iterative process beginning with independent reading and immersion in the data followed by identification of recurring themes, coding and categorising of the data.

Initial coding was undertaken inductively, with themes derived from the data to identify factors influencing treatment acceptance and non-acceptance. These themes were then deductively coded to the TDF^9,10^. MA (an applied health economist with qualitative training) independently coded an initial set of six transcripts, and one round of double coding (4% of the sample) was undertaken by EC (a behavioural scientist). Coding was compared and agreed upon between the coders, and any discrepancies were resolved by discussion to develop a preliminary coding framework.

The remaining transcripts were then independently coded by MA, with further rounds of iteration of the coding framework between MA, RG and EC. An iterative process was used where themes were discussed between members of the research team and modified accordingly (e.g. by merging / splitting existing themes, creating new themes / sub-themes or re-conceptualising themes). Themes were then grouped into high-level themes within each TDF domain and classified as either a barrier, an enabler, or a mixed influence on decisions to accept or decline preventive treatment. A narrative summary of findings was reported for each domain, with key domains identified based on the number of themes, the number of participants contributing to each domain, the degree of participant elaboration, and divergence or convergence of views across participants^13^.

The high-level themes were then grouped into cross-cutting themes across domains and deductively mapped into the COM-B model to characterise the Capability, Opportunity and Motivation factors underpinning decisions to accept or decline preventive treatment for TB infection^9,10^.

### Patient and public involvement

In collaboration with TB Alert (the UK’s national TB charity; https://www.tbalert.org/), a group of six PPI representatives was assembled, including people with lived experience of: TB disease; preventive treatment; being a carer for children with TB disease and TB infection; and working in migrant communities. The six-person patient advisory group provided input into recruitment procedures and the interview guide, and supported the original funding application.

## Results

### Overview of participants

A total of 25 participants were recruited and included in the analysis (Table 1). Median age was 34 (interquartile range 27 to 49) and most were male (16/25; 64%). The vast majority were born outside the UK (24/25; 96%%) and were either of Asian (n=10) or Black (n=14) ethnicity. Participants were mostly recruited through occupational health (n=14), migrant (n=6) or contact screening (n=4). Interviews were conducted between 10/06/2025 and 15/09/2025, lasted between 15 and 48 minutes, and were mostly conducted in person (23/25). A total of 14/25 (56%) of participants had accepted preventive treatment, with the remainder declining (n=7) or still being undecided (n=4). Thematic saturation was reached between the 20th and the 25th interview, with no new themes coded. All participants completed the interview, and none withdrew their data.

**Table 1:** Baseline characteristics of included participants. Participant demographic characteristics and preventive treatment status.

| Characteristic | N = 25<sup>1</sup> |
| --- | --- |
| Age | 34 (27, 49) |
| Sex |  |
| Female | 9 (36%) |
| Male | 16 (64%) |
| Ethnicity |  |
| Asian or Asian British | 10 (40%) |
| Black or Black British | 14 (56%) |
| White | 1 (4.0%) |
| Screening group |  |
| Migrant screening | 6 (24%) |
| Occupational health screening | 14 (56%) |
| Pre-immunosuppression screening | 1 (4.0%) |
| Recent TB contact | 4 (16%) |
| Preventive treatment |  |
| No | 7 (28%) |
| Undecided | 4 (16%) |
| Yes | 14 (56%) |
| <sup>1</sup>Median (IQR); n (%) |  |

### Barriers and enablers to acceptance of TB preventive treatment

Factors influencing participants’ decisions to accept or decline treatment for TB infection were mapped across 12 of the 14 TDF domains (Table 2, Figure 1). No factor was mapped into the domains of ‘Skills’ or ‘Reinforcement’. Table 2 presents the high-level themes generated within each domain, along with supporting quotes. Supplementary Table 1 provides a full list of themes and subthemes within each domain. The sections below provide a narrative summary of the 12 domains and the mapped high-level themes.

**Figure 1:**
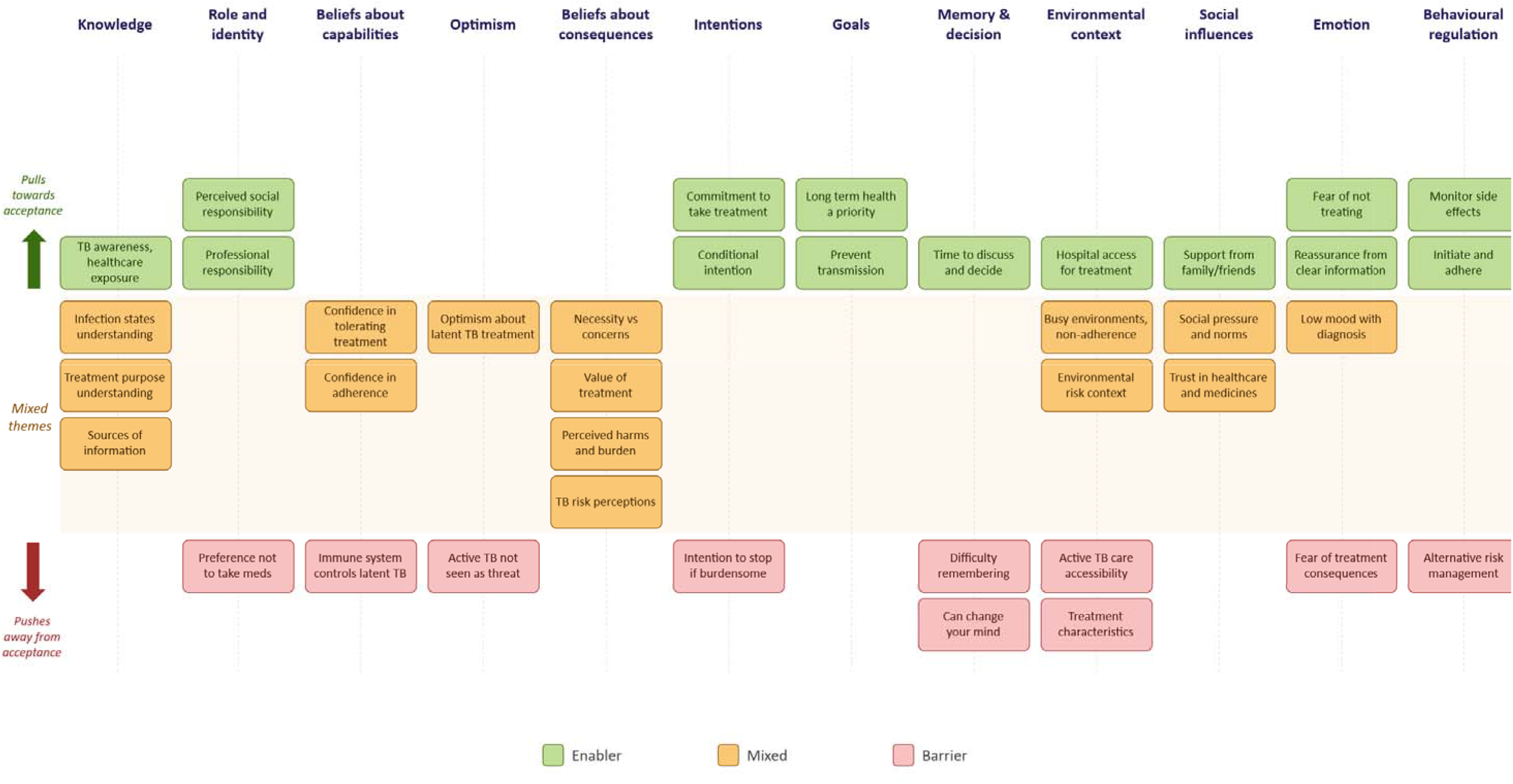
Overview of factors influencing acceptance of TB preventive treatment, mapped to Theoretical Domains Framework. Summary of barriers, enablers, and mixed influences on preventive treatment acceptance across Theoretical Domains Framework domains.

**Table 2:**
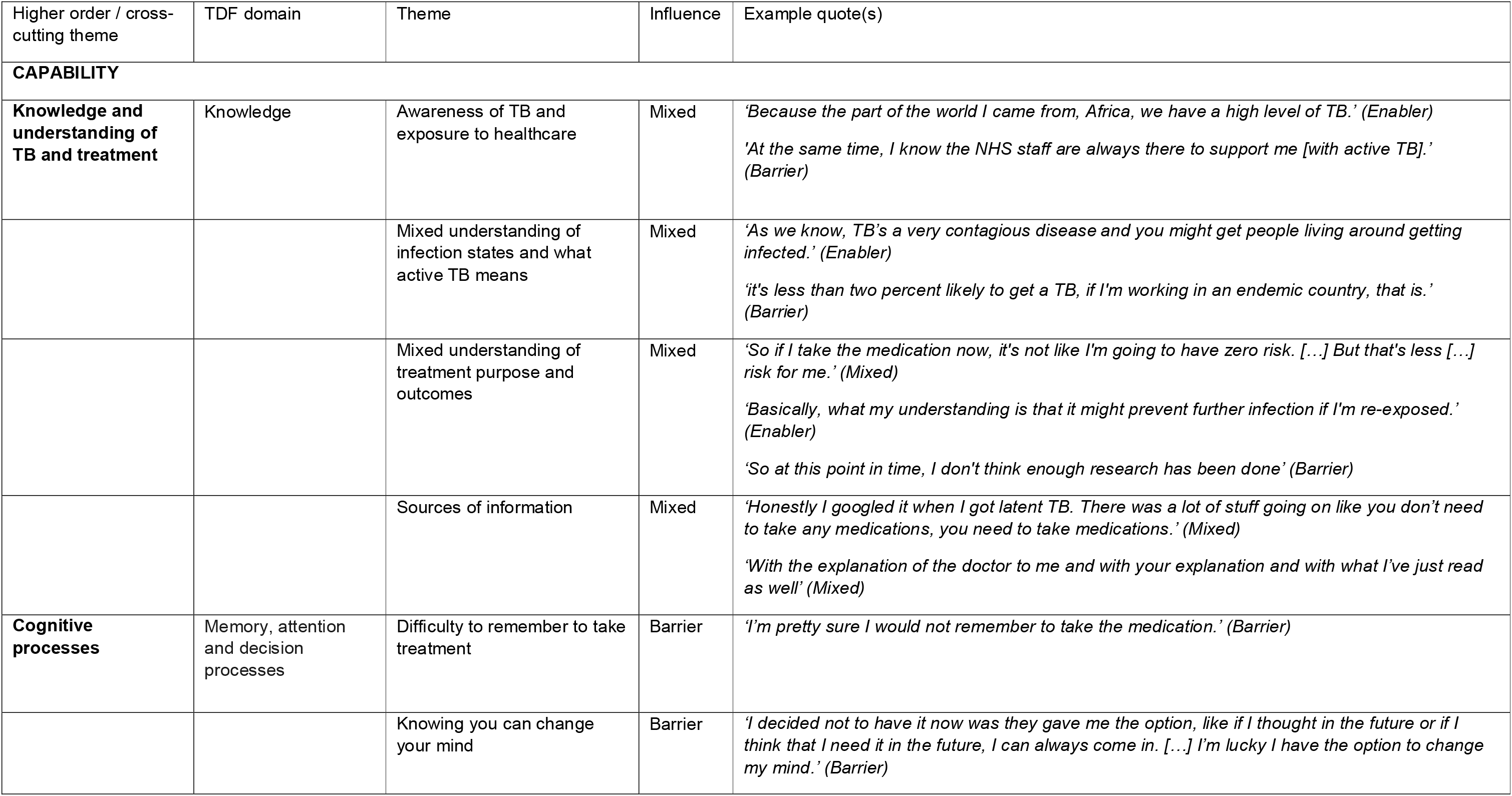
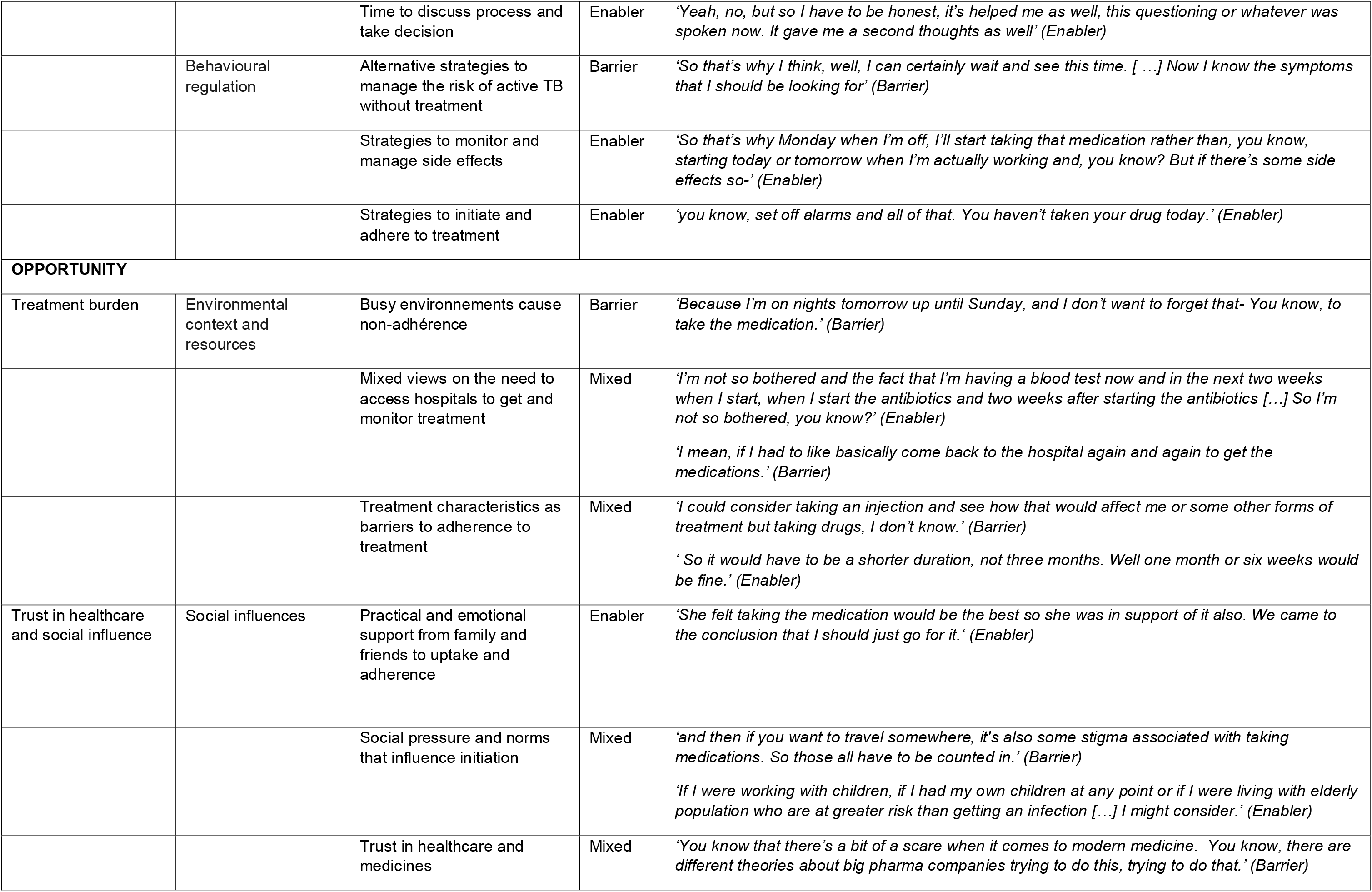
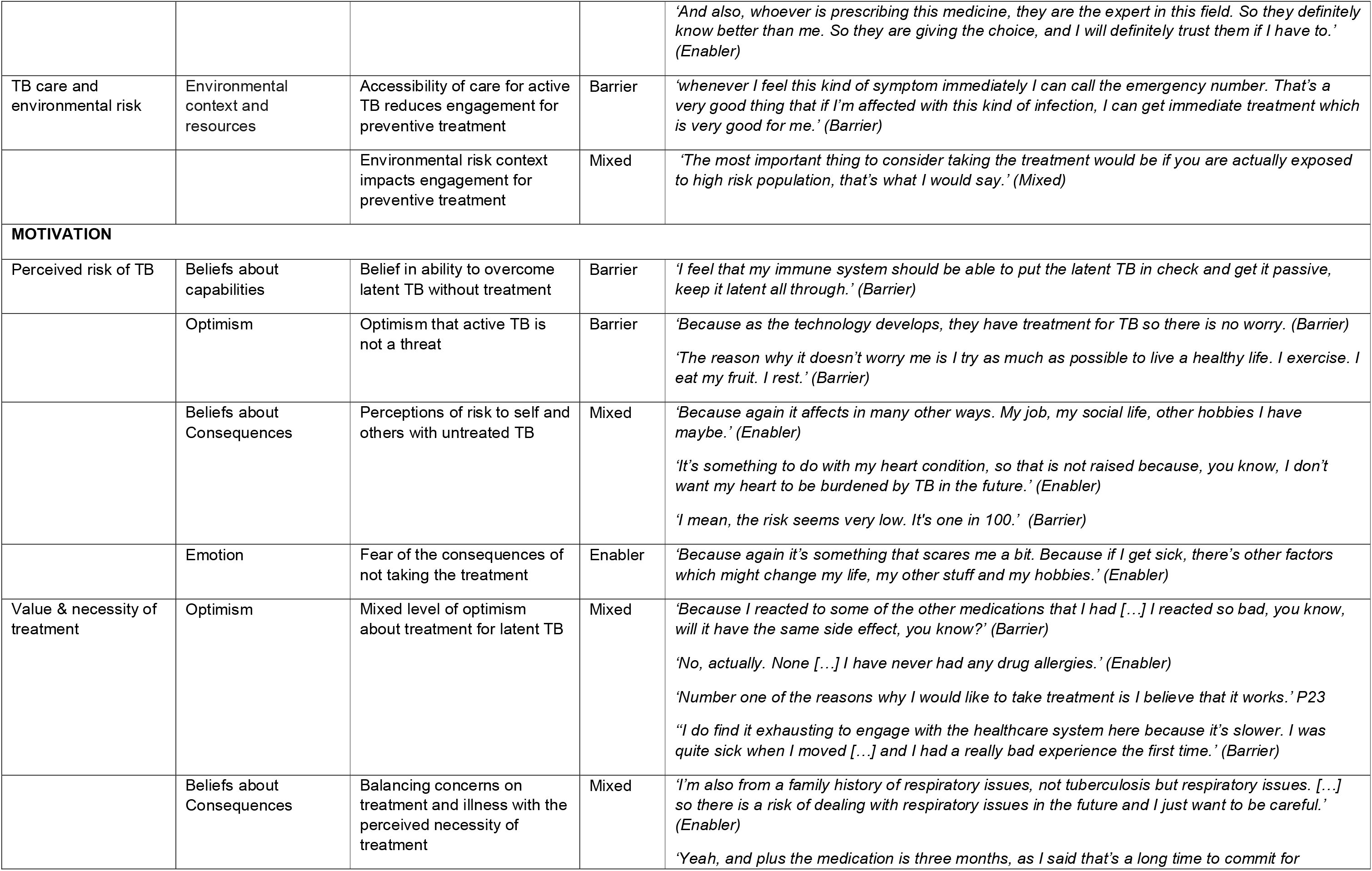
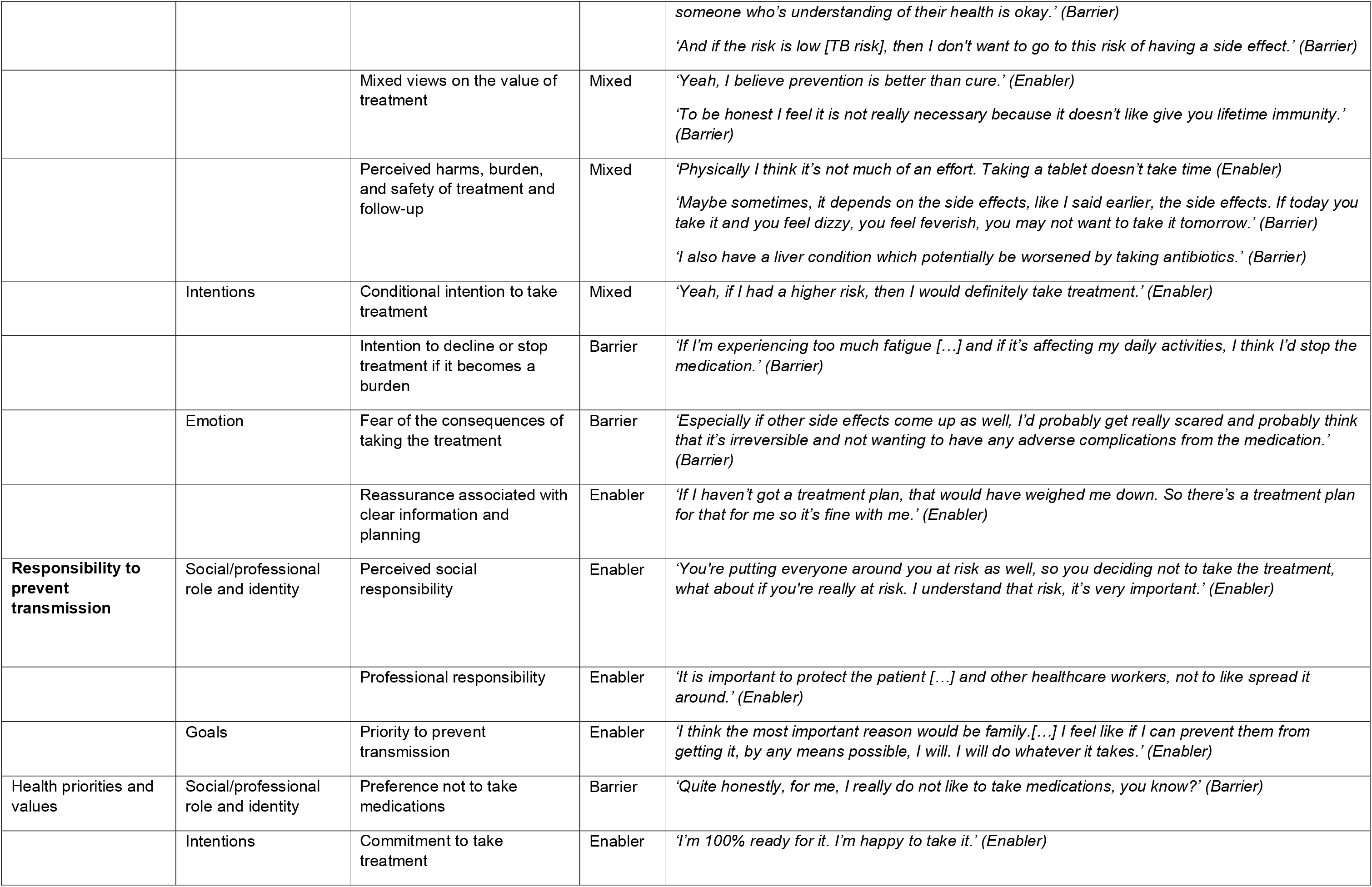
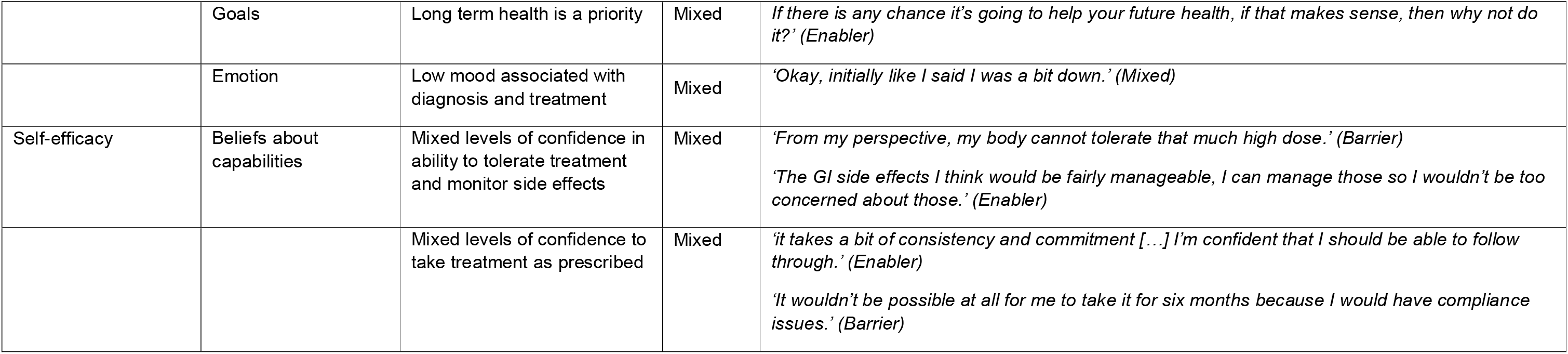
Barriers and enablers to acceptance of TB preventive treatment mapped to the Theoretical Domains Framework (TDF) and Capability, Opportunity and Motivation model of Behaviour (COM-B) domains. Themes, direction of influence on treatment acceptance, cross-cutting themes, and illustrative participant quotations mapped to TDF domains. Full TDF themes are shown in Supplementary Table 1.

### Knowledge

*Knowledge* had a mixed influence on decisions to accept or decline preventive treatment. Participants reported mixed understanding of TB infection states and of the purpose and outcomes of treatment. While participants were generally aware that preventive treatment reduces the risk of active TB, misconceptions that treatment protects against future re-exposure rather than current TB infection were reported. On the other hand, awareness that treatment does not provide lifetime immunity and a lack of knowledge about the evidence for preventive treatment sometimes acted as barriers. Knowledge of side effects and treatment burden had a mixed influence on preventive treatment acceptance. Participants’ awareness of the broader context, including environmental risk factors, familiarity with the healthcare system, and the availability of NHS support in the event of TB becoming active, also had a mixed impact on their willingness to accept the treatment. Different sources of information were mentioned, which can both enhance knowledge and lead to confusion.

### Social and professional role and identity

*Social and professional role and identity* had a positive influence on participants’ attitudes toward preventive treatment. Some participants reported a sense of moral or social duty to prevent harm to others and protect family, friends and the wider society. This sense of responsibility was particularly pronounced among some healthcare workers, viewing the decision to decline treatment as an unacceptable risk to patients and colleagues. Conversely, some participants expressed a personal aversion to medication, preferring to avoid medication unless they were experiencing symptoms or were seriously ill.

### Beliefs about capabilities

Most themes in *Beliefs about capabilities* had a mixed influence on treatment acceptance. Participants reported varying levels of confidence in their ability to tolerate preventive treatment and monitor side effects. Concerns from some participants included doubt about their physical ability to tolerate the treatment due to age or underlying health conditions, while others felt the treatment and its side effects would be easily manageable. Similarly, confidence in adherence varied. While some expressed strong commitment to following the prescribed regimen, others anticipated difficulties due to compliance issues or the physical properties of the medication itself. A notable barrier to treatment uptake emerged from some participants believing in their ability to overcome TB infection without treatment.

### Optimism

A common barrier within *Optimism* was participants’ positive outlook that active TB would be manageable or unlikely to pose a major threat. This was supported by confidence in treatment for active TB, and optimism about the positive impact of lifestyle changes in avoiding active disease. Participants’ broader optimism about the preventive treatment and its benefits was shaped by other motivational factors such as prior experiences with medication tolerance, treatment side effects, and healthcare (beliefs about consequences).

### Beliefs about consequences

*Beliefs about Consequences* emerged as a key domain, with a large number of themes developed, contributions and extensive elaborations from all participants and notable divergence in views. This domain had a mixed influence on preventive treatment acceptance. Participants weighed their perceived future risk of TB against their current health status, including the presence or absence of symptoms, age, comorbidities, family history, and perceived ability to cope with illness. These were balanced against the anticipated side effects, treatment burden, monitoring demands, and concerns about drug interactions in the context of comorbidities, all of which acted as barriers to uptake. Treatment duration was a particularly common concern, with some participants viewing a three-month regimen as too great a commitment, especially when the risk of TB was perceived as low. Others considered the treatment length manageable, or that even a small risk would justify preventive action regardless of the treatment burden. Anticipated consequences of progression to active disease generally motivated treatment acceptance, including personal harm, concerns that TB could worsen or complicate future healthcare for comorbidities, and the risk of transmission and burden to others.

Overall, participants had mixed views on the value of treatment. While some participants believed in a ‘prevention over cure’ approach and in the necessity to protect others from TB, others questioned the usefulness of preventive treatment. These concerns were driven by perceived limitations in protection, the likelihood of reinfection, uncertainty regarding treatment outcomes and perceived lack of evidence supporting the need for preventive treatment. However, among participants who shared these concerns, a few expressed potential regret if TB became active without treatment when deciding whether to accept the treatment.

### Intentions

*Intentions* toward preventive treatment varied across participants. Some participants expressed a firm intention to take preventive treatment, including maintaining commitment despite uncertainty about treatment effectiveness or changes in future TB risk. Others expressed conditional willingness to take preventive treatment, depending on factors such as follow-ups and testing or an increase in perceived TB risk to themselves or others. Conversely, some participants expressed intentions to decline or discontinue treatment if it became too burdensome, particularly if side effects were to disrupt daily activities or routines.

### Goals

Themes within the *Goals* domain had a positive influence on attitudes toward preventive treatment. Participants were motivated by a commitment to their long-term health, including protecting future wellbeing and preventing progression to active TB, particularly in the context of existing or anticipated comorbidities. A desire to prevent transmission to others, including family members and the wider community (a social influence), had an impact on their goals surrounding treatment acceptance.

### Memory, attention and decision processes

*Memory, attention and decision processes* acted predominantly as barriers to engagement with preventive treatment. Participants anticipated challenges in remembering to take preventive treatment consistently. Some also noted that their decision was not fixed and could be revised if their views or circumstances were to change. Others expressed a need for more time to discuss their options and reflect before reaching a final decision.

### Environmental context and resources

*Environmental context and resources* had a mixed influence on participants’ acceptance of treatment. The accessibility of care for active TB reduced motivation for preventive treatment among some participants, who felt reassured that symptoms could be managed if TB became active. Busy or demanding environments, including urban settings, eating patterns, lifestyle demands, and work constraints, were described as barriers to adherence. Environmental factors perceived to increase the risk of exposure or reinfection had a mixed impact. While some participants felt these increased other aspects of their motivation such as the need for preventive treatment (beliefs about consequences), others questioned its value given the likelihood of reinfection.

Participants expressed mixed views about hospital-based care. Some found follow-up visits and tests reassuring, while others discussed a lack of timely or local medical support. They also mentioned challenges in commuting to the hospital, seeing multiple hospital visits as a barrier to engagement. Participants described features of the treatment itself as barriers to engagement, including dose frequency, medication properties, side effects, and treatment duration. Longer treatment duration increased the perceived burden and may lower uptake. Participants also discussed how the frequency of doses, such as daily doses, could reduce adherence and uptake.

### Social influences

*Social influences* had a mixed influence on acceptance of preventive treatment. Practical and emotional support from family and friends facilitated both the decision to start preventive treatment and ongoing adherence. Participants described how social pressures and norms shaped decisions about starting preventive treatment, including anticipated stigma, the influence of others’ experiences of TB and its treatment, as well as interactions with vulnerable people. Trust in healthcare professionals, prescribed treatment, and the wider medical system generally supported engagement. However, reassurance that health professionals would be available to manage TB if it were to develop sometimes reduced motivation for preventive treatment.

### Emotion

*Emotion* influenced engagement with preventive treatment in both directions. Fear and worry about the potential consequences of not taking preventive treatment, including concern about developing active TB, missing symptoms, and living in high-risk environments, motivated some participants toward accepting the treatment. Conversely, fear and anxiety about the potential consequences of taking preventive treatment, including concerns about minor or serious side effects, the duration of treatment, taking too much medication, and possible effects on underlying conditions, acted as barriers. Some participants also described low mood and emotional distress related to TB diagnosis and the potential of taking preventive treatment. Reassurance was facilitated by clear explanations of TB infection and its defined treatment plan.

### Behavioural regulation

*Behavioural regulation* generally had a positive influence on engagement with preventive treatment. Participants described strategies to monitor and manage side effects and plan in advance to attend check-ups. Strategies to initiate and maintain treatment included fitting treatment around existing routines or creating new ones, using reminders, and timing treatment initiation to accommodate practical constraints. However, some participants described alternative approaches to managing TB without preventive treatment, including changing habits to reduce risk and self-monitoring for symptoms of active TB.

### Mapping to COM-B

High-level themes influencing decisions to accept or decline preventive treatment for TB infection were grouped into ten cross-cutting themes and mapped onto the three COM-B components, capability, opportunity, and motivation (Table 2, Figure 2). Under *capability*, the cross-cutting themes ‘Knowledge and understanding of TB and treatment’ and ‘Cognitive processes’ highlighted differences in psychological capability to engage with preventive treatment. Under *opportunity*, cross-cutting themes ‘Treatment burden’, ‘TB care and environmental risk’, and ‘Trust in healthcare and social influence’ reflected the external physical and social factors that shape decisions on whether to accept or refuse treatment for TB infection. Under *motivation*, the themes ‘Responsibility to prevent transmission’ and ‘Health priorities and values’ contributed to automatic motivation to treatment uptake, while ‘Perceived risk of TB’, ‘Value and necessity of treatment’, and ‘Self-efficacy’ shaped reflective motivation.

**Figure 2:**
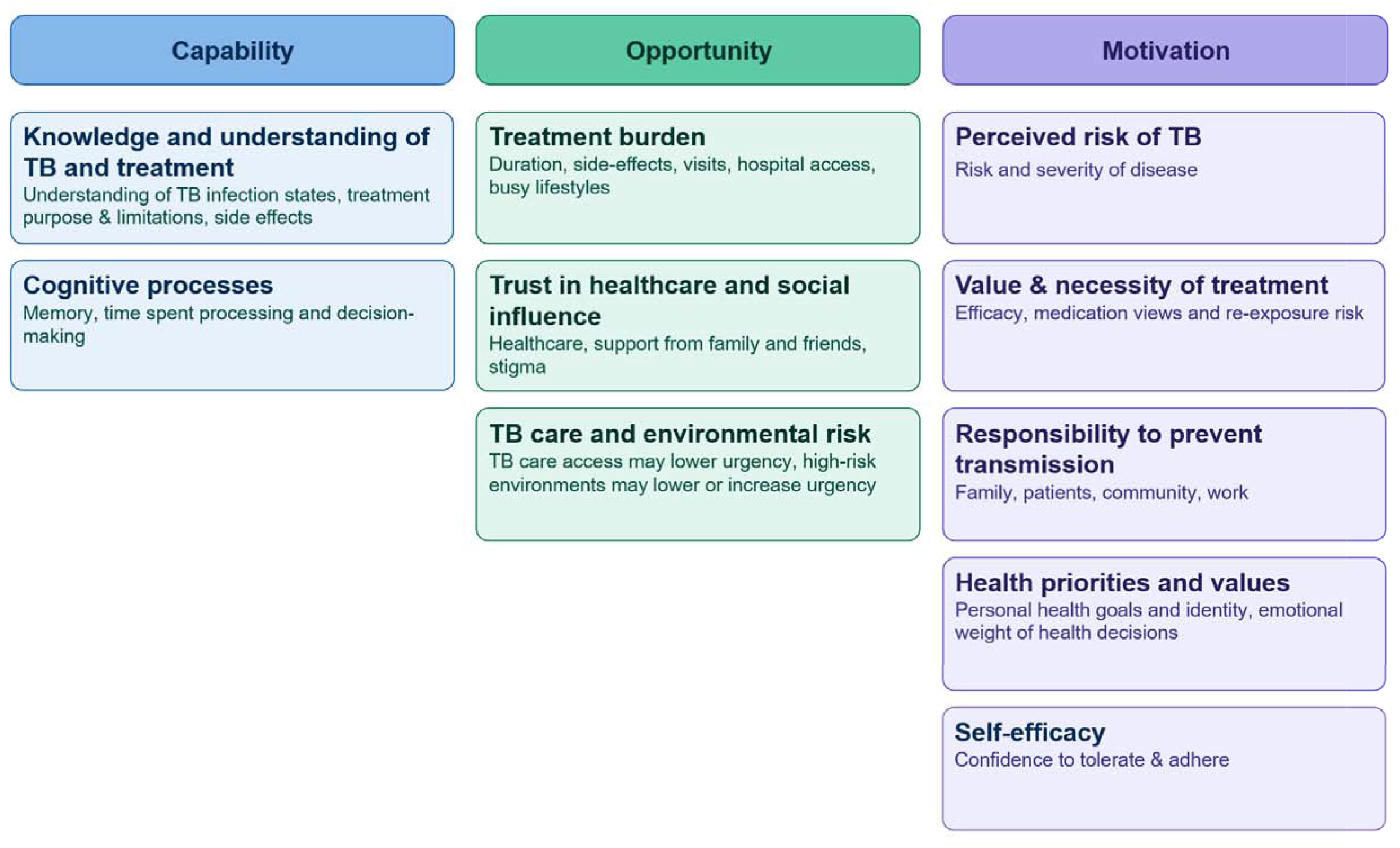
Overview of cross-cutting themes influencing acceptance of TB preventive treatment, mapped to Capability, Opportunity and Motivation mod Behaviour (COM-B) domains. Cross-cutting themes and mapping to Theoretical Domains Framework themes are shown in Table 2.

## Discussion

This study aimed to identify factors influencing participants’ decisions to accept or decline preventive treatment for TB infection. While prior studies have shown that knowledge is a key determinant of treatment acceptance^6–8^, using the TDF allowed us to systematically explore the range of individual, social, and environmental influences on this behaviour in the current study. Factors were mapped across 12 of the 14 TDF domains, reflecting the complexity of decision-making and treatment uptake in this context.

Participants’ knowledge of TB infection states, treatment purpose and outcomes, and broader contextual factors directly shaped their motivations such as perceived necessity of, and motivation to take, preventive treatment. For instance, awareness that treatment does not provide lifetime immunity, or a perceived lack of robust evidence supporting the need for intervention, reduced the motivation to accept treatment. While awareness of TB and worry about progression are commonly reported in the literature, our findings confirm that these do not necessarily translate into uptake^14^. Previous research has found a mixed influence of fear on treatment engagement; while fear of disease progression can motivate uptake, fear of side effects and treatment burden act as barriers^15,16^. These are consistent with our findings. Participants also identified anticipated stigma as a potential barrier to treatment, consistent with prior evidence that stigma can lower uptake^16–18^.

Participants’ confidence in their ability to tolerate and adhere to the medication influenced how they weighed the pros and cons of treatment. Those with low self-efficacy, whether due to anticipated memory challenges or perceived vulnerability related to age or underlying health conditions, often concluded that they were personally unable to complete the preventive treatment regimen, reducing the motivation to accept treatment regardless of clinical eligibility. Conversely, some showed high self-confidence in their own immunity, believing they could overcome TB infection without medication, which also reduced motivation to accept treatment. These findings illustrate how beliefs about capabilities shape intentions, which in turn influences treatment-related behaviour. While focusing on adherence rather than uptake, previous research has also suggested that self-efficacy and capacity shape outcome expectations and, in turn, influence motivation and medication adherence^19,20^.

External physical and social factors also played an important role in shaping motivational factors such as the perceived value of preventive treatment. Busy lifestyles, the hospital-based model of care, and characteristics of the medication itself increased the perceived burden of treatment and acted as barriers to uptake, consistent with evidence that treatment duration and side effects are reported barriers to preventive treatment for TB infection^14,17^. While our findings are consistent with the literature showing support from family and trust in healthcare professionals generally enhanced motivation, we also found that the availability of care for active TB paradoxically reduced the perceived necessity of prevention for some participants. Environmental and social factors therefore shaped the priorities that either reinforced or undermined engagement with preventive treatment.

Reasons for accepting treatment included protecting personal health, reducing future problems if current or future comorbidities develop, and preventing transmission to others. Some participants adopted a preventive approach regardless of their future TB risk. For those participants, a high future TB risk tended to reinforce uptake, but a low risk was perceived as ‘still a risk’. Other participants considered their risk of future TB too low to justify the treatment, with some also expressing optimism that good general health and lifestyle changes would prevent progression to active disease without treatment. However, some expressed willingness to reconsider their decision if their future TB risk were to increase. These findings reflect the role of necessity beliefs in treatment decision-making, and are consistent with evidence that low perceived personal risk of disease acts as a barrier to preventive treatment engagement^21,22^. Among participants who engaged with treatment, regulation strategies were important facilitators, such as integrating medication into existing routines, using reminders, and proactively planning for side effect management, in line with evidence that routine integration and reminder strategies support adherence across a range of conditions^17,23,24^.

A key strength of this study is the use of the TDF to systematically explore not only the range of factors influencing decisions around preventive treatment, but also their interconnectedness and the trade-offs participants made between competing barriers and facilitators. The study goes beyond individual-level factors to also capture the social and environmental influences that shape behaviour. The themes generated through this analysis provide a strong foundation for intervention development, as mapping barriers and facilitators to TDF domains helps explore how adherence challenges can be addressed, for example, by modifying external factors, such as treatment characteristics or care delivery, and by targeting internal factors, such as self-efficacy or knowledge. Furthermore, the TDF domains could be mapped onto the three components of the COM-B model, capability, opportunity, and motivation, which sits at the centre of the Behaviour Change Wheel and can support the evidence-based and theory driven selection of appropriate intervention strategies^25^. Another strength is the rigour of the analysis, which was supported by double coding of one interview and iterative refinement of the coding framework through discussion within the research team. These methods align with recognised quality standards for qualitative research and are frequently reported in health behaviour research^26,27^. Finally, the sample was representative of people offered preventive treatment in the UK and other low incidence settings, promoting generalisability. We included both participants who accepted and those who declined preventive treatment, thus enabling a comprehensive exploration of both facilitators and barriers.

A limitation of the current study is that we recruited from only two London sites; future studies could further examine the generalisability of our findings in settings with low and high TB incidence. The findings of the current study are being used to inform the design of a larger scale discrete choice experiment, to explore factors impacting treatment acceptance quantitatively in a wider population. Second, coding was done by a single researcher for most transcripts. This was mitigated by following guidelines and through iterative refinement of the coding framework within the research team, which included both experts in TB infection and a behavioural scientist familiar with the TDF^10^. Finally, social desirability bias may have influenced participants’ responses, particularly given that interviews were conducted in a clinical setting and with health professionals. Nevertheless, our analysis has identified in-depth facilitators and barriers to uptake and we included participants who declined treatment programmatically, suggesting the magnitude of this bias was likely small.

In summary, decisions to accept preventive TB treatment are complex and were driven by interacting capability, opportunity and motivation factors, rather than knowledge alone. Low perceived TB risk, high treatment burden and doubts about tolerability reduced uptake, while motivations to protect health and others supported engagement. These findings identify clear targets for intervention to improve shared decision making and acceptability- including improving risk communication, reducing burden and strengthening patient confidence.

## Data Availability

Interview transcripts cannot be shared due to risk of deductive disclosure; participant consent did not permit public data sharing. The full coding framework, themes, subthemes and illustrative quotations underpinning the analysis are provided in the supplementary materials.

## Footnotes

## Acknowledgements

The authors are grateful to all study participants and clinical staff who supported the study and contributed towards recruitment.

## Author contributions

RKG and EC conceived the study. MA, MN, LLR, EC and RKG contributed to protocol development. RKG, MN, RRK and SA led recruitment. RRK and SA conducted the interviews. MA and EC led the analysis, with input from RKG. MA, EC and RKG led manuscript writing. All authors reviewed the manuscript and contributed to interpretation.

## Funding

RKG is supported by the National Institute for Health Research (NIHR303184), Wellcome Trust (314897/Z/24/Z), the Royal Society (ICAO\R1\241109) and by NIHR Biomedical Research Funding to University College London Hospitals. MN supported by the Wellcome Trust (306550/Z/23/Z) and NIHR Biomedical Research Funding to University College London Hospitals.

## Declaration of interests

The authors have no competing interests to declare.

## Notes

### Competing Interest Statement

The authors have declared no competing interest.

### Clinical Trial

NCT07024836

### Author Declarations

The London - Brighton & Sussex Research Ethics Committee gave ethical approval for this work (25/LO/0125).

