## Supplementary Appendix for "Patient acceptance of preventive antibiotic treatment for tuberculosis: a qualitative study"

### Interview guide questions

#### General

1. Please can you tell me a bit about yourself and how you have come to be here today?

#### Treatment

1. If you have been diagnosed with latent TB infection, you may have been offered preventive antibiotic treatment - to reduce the chances of becoming unwell with TB in the future. What comes to your mind when you hear about treating latent TB with preventive antibiotics?

- Probe: What do you understand by “preventive” antibiotics?
- Probe: What are your views on preventive antibiotics?
- Probe: Do you think taking the antibiotics will help you to stay healthy?

2. Are you taking preventive antibiotic treatment for latent TB infection, or planning on starting this? Can you tell me more about this?

- Probe: How do you feel about taking preventive antibiotics?
- Probe: What are the things (factors) that made you want to take preventive antibiotics?
- Probe: What are the things (factors) that made you not want to take preventive antibiotics?
- Probe: Do you have any concerns about taking the treatment?
- Probe: How much time or effort do you think it would be to take the antibiotics?
- Probe: Do you think taking the treatment is necessary? If so, why?
- Probe: To what extent would taking your medication affect your daily routine? [Probe for differences between weekdays and weekends, during holidays.]
- Probe: What might make it difficult for you to take the treatment?
- Probe: Tell me about any times that you might be more or less likely to miss a dose?
- Probe: Describe to me any thoughts about what might make it easier for you to take the treatment?

3. How did you decide whether or not to take the treatment?

- Probe: How did/do you weigh the good things about taking treatment versus the bad things that made/make you not want to it?
- Probe: Did/will you talk to anyone about taking the treatment?
- Probe: If the treatment lasted longer, for example six months instead of three, how would that change the way you feel about taking it?
- Probe: If the treatment required check-ups or tests every two weeks during the three months, how would that influence your view of it?

4. If you have already started taking treatment, how have you found it?

- Probe: What information did you receive about the treatment before starting?
- Probe: What were your initial thoughts and expectations of the treatment before you started it?
- Probe: Then after starting, how did it go/is it going?
- Probe: What would you advise someone who starts the same treatment that you have had?

5. What do you know about the side effects of treatment for latent TB?
6. When the treatment was offered to you, did you discuss potential side effects with the doctor? How did you feel about what you were told?

- If you have already started taking treatment, do you experience any side effects due to treatment? If yes, could you describe them [Probe: frequency, severity, how do you manage them]
- Are you anxious about side effects potentially occurring? [Probe: what concerns you the most?]
- How would these side effects impact your daily life? [Probe: work, social life, sleep, mood, physical activity, relationships]
- Can you think of a situation where the experience of side effects could become a reason to stop or refuse the treatment?

7. Could you tell me about your understanding of latent TB infection and what it means to you?

- Probe: What do you understand when you hear latent TB infection?
- Probe: How do/did you feel about the idea of getting tested for latent TB infection?
- Probe: Do you understand what your test result means?
- Probe: How do you feel about your test result?

#### Future TB risk

1. What do you understand about your risk of becoming unwell with TB in the future?

- Probe: Does/did your future risk of TB worry you? If so, why?
- Probe: Does/did your future risk of TB make you want to take preventive antibiotics?
- Probe: If your future risk of TB (without any treatment) were to change, would this make you more or less likely to take preventive antibiotics? For example, if your risk of TB without treatment was higher (lower), how would you feel about this? Would it make it you more or less likely to take treatment?
- Imagine you took the preventive treatment now, but later tested positive with active TB, how would you react? Would that make you more or less likely to take preventive treatment now?

#### Concluding

1. What would you consider the most important reason for taking latent TB treatment?
2. What do you think is the biggest reason that stops people from taking latent TB treatment?
3. Is there anything else you would like to discuss, or do you have any questions?

#### Supplementary Table 1: Factors that positively or negatively influence the uptake of preventive treatment within each theoretical domain

Codes represent factors perceived to influence the choice to take the preventive treatment within each TDF domain. Initial codes are classified as either a positive, negative or mixed influence on treatment uptake.

| Domain | High-level Themes | Codes | Influence | Supporting quotes |
| --- | --- | --- | --- | --- |
| 1. Knowledge  (An awareness of the existence of something) | 1. Awareness of TB and exposure to healthcare  Participants’ awareness of their wider context and the healthcare system (e.g., environmental exposure risks, familiarity with healthcare, and the availability of NHS support if TB became active). | Knowing the NHS will support if active TB | - | *‘at the same time I know the NHS staff are always there to support me so there is no worry.’* P10 |
|  |  | Knowledge of environmental risk factors (workplace or country with a high transmission rate) | +/- | *‘If you check the statistics, you see that quite a lot of like- The TB prevalence in [home country] is a bit high.’* P02  *‘I work in [workplace] and pre-disposed, […] I’m working in a high-risk environment. […] You could be looking after someone with TB.’* P04 |
|  |  | Limited experience of illness and healthcare | - | *‘There’s a negative perception of NHS and I’ve come to realise that people, actually those people are the one that never been to hospital as yet […] So I think there’s an element of that in here that, you know, because they’ve never been unwell and so they, the opinion is that- It’s a very small percentage’. P04* |
|  | 2. Mixed understanding of infection states and what active TB means  Participants’ understanding of TB infection states (latent vs active), including awareness of the absence of symptoms in latent/early active TB, and awareness of active TB manifestations and infectiousness. | Knowing that current stage of TB is latent | - | *‘I was more about whether I have TB now. I mean, I wasn't sure. Am I suffering from TB now? […] Before I met the doctor, I didn't know that it was latent.’* P07 |
|  |  | Knowing that latent and early active TB have no symptoms | +/- | *‘and my current condition, I don't have any symptom.’* P07  *‘I’ve had contact with someone with TB and most of the time people with latent tuberculosis aren’t always symptomatic, they maybe don’t feel anything. Like it’s not active, it’s still dormant.’* P02 |
|  |  | Knowing that you can spread active TB | + | *‘As we know, TB’s a very contagious disease and you might get people living around getting infected.’* P19 |
|  |  | Mixed understanding of latent TB infection | +/- | *‘So latent TB, what I understand is that I have the presence of the [s/l tuberculosis 00:15:04] bacteria in my body and I carry it around. It’s not really doing anything and they’re fairly inactive. There is the risk that they will spread and become an active TB infection.’* P15  *‘from all the results and then all the interventions that I have received so far, it's less than two percent likely to get a TB, if I'm working in an endemic country, that is.’* P11 |
|  |  | Understanding of active TB and its manifestations | +/- | *‘because of my background, like I said earlier, I understand how it can be once it becomes active […] I wouldn’t want something of such’.* P02  *‘What I know about TB is it’s an airborne disease. So it usually affects your respiratory system. So if it’s not treated early, it can be fatal.’* P22 |
|  | 3. Mixed understanding of treatment purpose and outcomes  Participants’ understanding of the purpose and expected outcomes of preventive treatment, including awareness that treatment does not provide immunity. | Clinical knowledge of preventive treatment | +/- | *‘As I said, I also come from the nursing background so I definitely know what are the side effects and indications. […] on the basis of my past experience with my clinical placement and all these things, I choose to not take.’* P10  *‘So the idea of treating or certainly preventative treatment is a straightforward one for me because of my background.’* P02 |
|  |  | Knowing that preventive treatment does not give you immunity | - | *‘I was told that it will not- I think the key thing that I got from the information is it is not, it would not like confer a lifetime immunity, even if I take the medication, there are chances if I get exposed to TB I might still get it again’.* P05 |
|  |  | Knowing that side effects are minor | + | *‘I know that they tend to occur within the first week or two and then they settle down. I know that for the most part they’re relatively not serious or minor. It’s mostly GI upset and abdominal symptoms.’* P14 |
|  |  | Knowing that the risk of serious side effects is low | + | *‘But in as much as there is no health like a major or severe complication […] the consultant mentioned that there could be but in a very rare occasion.’* P02 |
|  |  | Lack of knowledge (evidence gaps) about clinical need for treatment | - | *‘Is there any other blood test that can be done really to determine the proportion or the percentage which I could have used as a benchmark to like, okay, so latent but now it’s 90% progression or 70%. […] Now I know it’s latent but at what stage of latency is it? Is it really progressing, is not, he’s carrying or whatever. I don’t know.’* P01  *‘It might not be in the future. It might be considered that the concept of latent TB or the concept of preventative antibiotics are all just going to be absolute in maybe one or two years or in five years. So at this point in time, I don't think enough research has been done.’* P11 |
|  |  | Mixed understanding of treatment outcomes | +/- | *‘So if I take the medication now, it's not like I'm going to have zero risk. […] But if I take the medication, that's less, less, less risk for me.’* P18  *‘Basically, what my understanding is that it might prevent further infection if I'm re-exposed. That is my basic understanding.’* P11 |
|  | 4. Sources of information  The information sources participants used to build their understanding of TB infection and preventive treatment, including health professionals and online resources. | Health professionals | +/- | *‘Yeah, with the explanation of the doctor to me and with your explanation […] more explanation like this so I see a positive side to it.‘* P12 |
|  |  | Online resources | +/- | *‘I actually go online to check, when I got the diagnosis in the first- I went online to check. I got some, you know, some- I went on Twitter, I went on the social media and all of that.’* P02 |
| 3. Social/professional role and identity | 1. Perceived social responsibility  A sense of moral or social duty to prevent harm to others and protect family, friends and society. | | + | *‘You're putting everyone around you at risk as well, so you deciding not to take the treatment, what about if you're really at risk. I understand that risk, it’s very important’.* P01 |
|  | 2. Preference not to take medications  A personal preference of avoiding medicines, or only taking medication when symptomatic or seriously ill. | | - | *‘Quite honestly, for me, I really do not like to take medications, you know?’* P02 |
|  | 3. Professional responsibility  Participants’ sense of obligation linked to their work or professional role, such as taking care of others or limiting risks. | | + | *‘Basically, as a healthcare worker […] Definitely you want to care for people, you want to do a lot of things for them and all, and you don’t want to care for them and at the same time put them at risk.’* P02  *‘It is important to protect the patient, not to- And other healthcare workers, not to like spread it around.’* P05 |
| 4. Beliefs about capabilities | 1. Belief in ability to overcome latent TB without treatment  Participants’ confidence that latent TB could be managed without preventive treatment, often reflecting confidence in their own immune system. | | - | *‘I feel like when you get told it’s latent, you're like, oh, I can fight it. It's latent. It's not going to show up. I'm fine.’* P06  *‘I feel that my immune system should be able to put the latent TB in check and get it passive, keep it latent all through.’* P19 |
|  | 2. Mixed levels of confidence in ability to tolerate treatment and monitor side effects  Participants reported mixed confidence in their ability to tolerate treatment and manage side effects, with some expressing concern that their body might struggle to adapt to the medication dose or its effects. | Confidence in ability to monitor and cope with side effects | + | *‘The GI side effects I think would be fairly manageable, I can manage those so I wouldn’t be too concerned about those.’* P14  *‘so if anything goes wrong with me, I can pick it up. So let’s assume then I start taking these and I start noticing some things I know is from the medication so I can easily report it or stop it, that’s it.’* P12  *‘From my perspective, my body cannot tolerate that much high dose.’* P10 |
|  |  | Uncertainty about ability to adapt to treatment and side effects | - | *‘My concern is my age, if my body is not able to fight it. Maybe it will lead to other complications.’* P20 |
|  | 3. Mixed levels of confidence to take treatment as prescribed  Participants varied in their confidence in being able to adhere to the prescribed treatment, with some reporting that they usually find it challenging to take medication regularly or might physically struggle with aspects of the medication, such as swallowing tablets. | Confidence in ability to take treatment as prescribed | + | *‘Yeah. Yeah, I think I can do that, yeah. Once I make a certain decision, yeah, why not?’.* P05  *‘So for this I’m going to, you know, it takes a bit of consistency and commitment and I’m quite- I’m confident that I should be able to follow through, you know, should be able to’* P02 |
|  |  | Lack of confidence in their ability to adhere to treatment | - | *‘It wouldn’t be possible at all for me to take it for six months because I would have compliance issues.’* P16  *‘But I think it’s mental practice, and I’m not the best at taking medication. I don’t take my vitamins as well so I think objectively it’s not really difficult but I’m not [unclear 00:06:22] so I’m not sure how that will work.’* P13  *‘The size of the tablets I guess if they’re difficult to swallow, it would make it quite difficult.’* P20 |
| 5. Optimism | 1. Mixed level of optimism about treatment for latent TB  Participants' varying levels of optimism about treatment and its benefits, shaped by past experiences with medication tolerance, side effects, and healthcare. | Confidence and positivity about treatment | + | *‘I'm fine with it actually. There might be some side effects or something in the beginning, like maybe stomach aches. But that’s something you get over pretty quickly. There’s nothing to be scared about and I'm fine with it.’* P8  *‘I should be okay. I'm looking forward for it to work anyway. […] Being on antibiotics for three months, but I'm just being positive that I will not develop side effects. ’* P17 |
|  |  | Optimism based on past experience of tolerating medicine well | + | [Do you have anything else that you think might make it difficult to carry on and take this treatment?] *‘No, actually. None […] I have never had any drug allergies.’* P05 |
|  |  | Optimism that the treatment will help to stay healthy | + | *‘These are different because the paracetamol like would just- It’s more like comparing if you follow that, but this is much more structured towards, you know, helping clear off the latent tuberculosis, you know?’* P02  *‘The reason is for me to be healthy by having it’.* P03 |
|  |  | Pessimism based on past experience of side effects | - | *‘Because I reacted to some of the other medications that I had, and it’s quite a bad reaction, like muscles at the start, I reacted so bad, you know, will it have the same side effect, you know?’* P04 |
|  |  | Pessimism based on past experience with the health care system | - | *‘I do find it exhausting to engage with the healthcare system here because it’s slower. I was quite sick when I moved [unclear 00:25:36] and I had a really bad experience the first time.’* P13 |
|  | 2. Optimism that active TB is not a threat  Participants' positive outlook that active TB would be manageable or not pose a major threat, including confidence in treatment for active TB, and optimism about the positive impact of lifestyle changes. | Confidence in treatment for active TB | - | *‘Because as the technology develops, they have treatment for TB so there is no worry. I can still do my life better with the antibiotic treatment.’*  P10 |
|  |  | Optimism about health or lifestyle changes | - | *‘No, not now because as I told you, I changed so many dietary, I do my dietary changes and turning my life towards a healthy life so there is no worry, yeah.’* P10  *‘The reason why it doesn’t worry me is I try as much as possible to live a healthy life. I exercise. I eat my fruit. I rest.’* P19 |
|  |  | Positive about having latent TB and not active TB | + | *‘When they explained that I was latent, I was much more happier.’* P07 |
| 6. Beliefs about Consequences (Acceptance of the truth, reality, or validity about outcomes of a behaviour in a given situation) | 1. Balancing concerns on treatment and illness with the perceived necessity of treatment  Participants weighed their perceived future risk of TB against their current health status (symptoms or ability to cope with illness), including the presence or absence of symptoms, age, comorbidities, family history, and perceived ability to cope with illness. Participants weighed their perceived future TB risk and current health status against the anticipated treatment burden and the risk of side effects. | Anticipated increase in risk of TB if comorbidities worsen | + | *‘So I’m worried that my heart will at some point become more unwell and that puts my sort of like immune system a bit more weaker, and then when you have a dormant infection that they can deal- Opportunistic point for them to become enacted, and this difficult too.’* P04 |
|  |  | Belief that age influences future risk of TB | + | *‘Although for my age bracket, according to the statistics that I saw around 5% actually like- Like 5% of people in my age bracket in five years are likely to, you know, [s/l gravitate 00:13:31] from the latent stage to the active stage’.* P02  *‘The latent is that I might have contracted it when I was young or been unaware. […] So I’m willing to take a risk because if it was at a very young age, I’m [age], that means my chances of not activate are higher than it being activated so I’m willing to take the risk.‘* P20 |
|  |  | Concerns about treatment duration | - | *‘I’d feel bad, I might feel somehow because probably I have settled my mind to three months, then if it was to be increased along the line, it will affect my thinking.’* P15  *‘Yeah, that’s exactly what I felt, you know, and the volume of the treatment is quite worrying to be honest, you know?’* P01 |
|  |  | Perceived increased necessity of preventive treatment due to comorbidities or family history | + | *‘I’m also from a family history of respiratory issues, not tuberculosis but respiratory issues. […] so there is a risk of dealing with respiratory issues in the future and I just want to be careful.’* P13 |
|  |  | Perceived lack of necessity as no symptoms | - | *‘If you're not having the condition and it’s proven that the virus is not active, why should I go on.’* P01  *‘why am I taking medication for something that I don’t have any symptoms for.’* P05 |
|  |  | Weighing future risk of TB and current health status | - | *‘Because of what I did tell the doctor or the consultant, I thought the chances of the infection getting worse would be less than more likely and that being 1% and my decision was based on that and my age, my lifestyle, my habits, my behaviour [unclear 00:20:03].’* P20 |
|  |  | Weighing current symptoms and treatment burden | - | *‘Yeah, and plus the medication is three months, as I said that’s a long time to commit for someone who’s understanding of their health is okay.’* P04 |
|  |  | Weighing future risk of TB and treatment burden | - | *‘I just feel that for me personally, even though I understand there is a risk the TB could reactivate, I don’t feel that that risk justifies taking three months of antibiotics.’* P14 |
|  |  | Weighing present health status vs risk of side effects | - | *‘Like I mentioned, you know, side effects basically and then you’ll end up having problems that you would need to start taking time off work and, you know, basically feeling unwell. Something that you were perfectly fine and then you started taking the antibiotics and then you end up opening something else entirely. So that was just a concern.’* P05 |
|  |  | Weighing risk of active TB vs risk of side effects | - | *‘And if the risk is low, then I don't want to go to this risk of having a side effect.’* P07 |
|  | 2. Mixed views on the value of treatment  Participants weighed the perceived value of preventive treatment against concerns about side effects, uncertainty about treatment outcomes and perceived limitations in protection.  Participants’ varying beliefs about the usefulness and benefits of preventive treatment regardless of the TB risk. | Acceptance of potential ineffectiveness of treatment or side effects | +/- | *‘When I heard about the side effects, fever, tiredness and all that but it doesn’t matter.’* P09  *‘I’ve asked about the side effect […] which to me, I said okay, it’s about taking the risk and if there is any reaction’* P12 |
|  |  | Belief in benefit of treatment even if reinfection was likely | + | *‘ you were to take the antibiotics now like you’re doing, got rid of the bacteria and then you caught it again in the future, how would that make you feel?*  *RES: I would believe that I’m more prone to it so I will still go back for treatment.’* P12 |
|  |  | Belief in benefits of preventive treatment despite burden | + | *‘It depends on the risk of the infection. If it was something that I know yes, it will be needed, it shouldn’t be anything for me to worry, even if it is to come every three days, I would know I have to.’* P15 |
|  |  | Belief in benefits of preventive treatment regardless of risk | + | *‘I don’t want any risk, even though it’s low, as low as- All I want is it’s going to be prevented’. P03*  *‘I think it’s the, you know, the chance of getting TB, I think that worries you, whether it’s 1% or 10%.’* P04  *‘If it is higher, definitely to push me towards taking the medication. But however, if the chances reduce it might not really affect me taking the medication because like I said, it is not because of me coming down with the TB, it is how it’s likely going to impact future health problems. So that’s the reason why I’m taking it.’* P08 |
|  |  | Belief in prevention over cure | + | *‘Yeah, I believe prevention is better than cure.’* P13 |
|  |  | Belief in the benefits of preventing TB at a young age | + | *‘Get it sorted at a young age with less responsibilities, more active and I can manage it much better.’* P06 |
|  |  | Belief that check-ups provide clinical information to understand treatment effectiveness/side effects | + | *‘No. I feel the check is just to see if there’s improvement so it’s fine. If you are not checked, you won’t know if there’s improvements or not. […] So if you are checked you will know okay there is improvements in the medication you are taking, you can just carry along so it’s fine.’* P16  *‘I would still have to come so that they can do some tests for me to be sure that the medication I'm taking does not trigger another thing which we both agreed on. So the way he actually explained it to me, I'm more convinced to take my medication.’* P23 |
|  |  | Belief that reinfection is likely due to ongoing exposure | +/- | *‘I’m thinking if I finish my … training, I might want to go back home and it’s quite an endemic zone there, so I might get re-infect again, so what is the point of taking the antibiotics?’* P05  *‘Because I’m still working in [workplace] and within an infectious- So I mean, who knows, it might be dormant now, you know, I’m always exposed to this. So there’s an element of that.’* P04 |
|  |  | Belief that the treatment will keep others safe | + | *‘So having the treatment will be an advantage for the environment as well, not to spread by me. Keep myself safe as well and people around me to be safe as well.’* P12 |
|  |  | Belief that treatment will help to stay healthy/prevent disease | + | *‘I don’t want to have that disease, that’s why I’m taking those antibiotics, just to be sure’.* P03 |
|  |  | Lack of belief in the effectiveness of preventive treatment | - | *‘Because prophylactic antibiotics have shown that they don't actually work in all the cases. So I'm not that convinced that antibiotic will always keep you healthy or prevent any infection taken prior to the infection.’* P11 |
|  |  | Perceived limited benefit if no immunity provided by treatment | - | *‘To be honest I feel it is not really necessary because it doesn’t like give you lifetime immunity, and because you can still get in contact with TB […] I don’t see the point actually if you can be re-infected later on’* P05 |
|  |  | Perceived low benefit from treatment due to ongoing exposure to TB | - | *‘I might want to go back home and it’s quite an endemic zone there, so I might get re-infect again, so what is the point of taking the antibiotics?’* P05 |
|  |  | Perceived uncertainty of treatment outcome | - | *‘Another thing is even if you take the medication, there is no test that you would take to ensure that, okay, now the- You don’t have the antibodies anymore, it is wiped off completely out of your system, do you get what I mean? So that’s another thing that I’m like, why am I even taking it?’* P05 |
|  |  | Weighing risk of side effects vs future benefits | +/- | *‘Well like I said earlier, because you don’t know what is going to happen in the future, you know? […] But then the chances of having the side effects is quite minimal so- […] Exactly, yeah, just weighing the risks, yeah’.* P05 |
|  | 3. Perceived harms, burden, and safety of treatment and follow-up  Participants’ beliefs about the potential harms and burden of treatment and follow-up, including side effects, treatment and monitoring demands, and added complexity or harm related to comorbidities. | Anticipated harm of treatment | - | *‘there are so many side effects. Sometimes it may be our bodies are not able to tolerate those antibiotics and also side effects definitely affect our kidney function, liver function.’* P10  *‘as I said, the body reacts differently to each drug. If I can’t accept it, it could probably then lead to other forms of medication being taken and then lead to other complications which I’m not willing to [unclear 00:21:27].’* P20 |
|  |  | Anticipated negative consequences of poor adherence | - | *‘I feel sometimes you may forget to take the medication and it can affect because it’s a daily medication and you have to take it.’* P09 |
|  |  | Belief that the preventive treatment is safe | + | *‘I think the main thing was hearing the side effects of the treatment and hearing that the side effects aren't something I should be too worried about.’* P06 |
|  |  | Beliefs that side effects may reduce adherence | - | *‘Maybe sometimes, it depends on the side effects, like I said earlier, the side effects. If today you take it and you feel dizzy, you feel feverish, you may not want to take it tomorrow.’* P16 |
|  |  | Perceived burden of follow up and check-ups | - | *‘it also affects my time schedule as in London people are very hurried for their schedules so yeah, it definitely affects my daily routine so that’s why.’* P10 |
|  |  | Perceived burden of treatment | +/- | *‘Yeah, because the time period’s too long. If it’s like a week or say two weeks or a month, I’d say, you know what- Three months and continuous treatment is only when there is no option’.* P01  *‘It’s quite long, three months’.* P04  ‘Yeah, and again taking medication for three months. It is such a commitment’. P05  *‘Physically I think it’s not much of an effort. Taking a tablet doesn’t take time*  *‘ P13*  *‘Just because I'm on my repro age, and I have a husband, so we're actually on our time of having baby. So this is the only concern I have, but probably three months just short time. So we can just do that after the treatment.* P18 |
|  |  | Perceived harm or treatment complexity due to comorbidities (drug interaction) | - | *‘No, actually. None, because I’m not taking any medication- And I don’t have any chronic illness and I’m not taking any other medication, so I’m not thinking of drug interactions.’* P05  *‘I also have a liver condition which potentially be worsened by taking antibiotics.’* P14 |
|  | 4. Perceptions of risk to self and others with untreated TB  Participants’ perceptions of the severity of their future risk of TB, and the perceived influence of environmental factors on the risk.  Participants’ beliefs about the potential consequences if TB progressed to active disease, including anticipated harm to themselves, such as concerns that TB could worsen or complicate future healthcare for comorbidities, risk of transmission and burden to others. | Anticipated burden on others if TB becomes active | + | *‘It will stop my family as well because they will want to look after me, they will want to know so it will stop so many things.’ P12* |
|  |  | Anticipated harm if TB becomes active | + | *‘Of course, because I don’t want to have that tuberculosis too, for me because that’s a bad thing in any person.’* P03  *‘I’ve seen someone with a worse case in [workplace], and with my current health it’s not a good combination and hence I’m open to take this medication.’* P04  *‘I feel like I cannot work as a nurse if I get the TB, which is will be difficult for me because that's the only sort of income that I can be, you know, could be alive.’* P18  *‘So I'm imagining in future what will happen to me. I’ll be coughing up blood before I get tested and know that I have TB and that might be too late for me.’* P22  *‘Because again it affects in many other ways. My job, my social life, other hobbies I have maybe.’* P09 |
|  |  | Anticipated risk of transmitting TB to others | + | *‘I don’t want my family to be contaminated as well with the disease.’* P03  *‘when it becomes active you become a risk to society.’* P02 |
|  |  | Belief that latent TB could worsen or complicate health care for potential future comorbidities | + | *‘so it is not the actual risk of developing TB later on, it is the possibility of having other comorbidities, if that makes sense? […] it might affect future health issues because it might limit your options for treatment later in life if by chance somebody comes down with a disease or something.’* P05  *‘to prevent symptoms coming up in the future and it hitting you when you're older or when you have a different type of illness.’* P06  *‘It’s something to do with my heart condition, so that is not raised because, you know, I don’t want my heart to be burdened by TB in the future.’* P04 |
|  |  | Concern about risk of TB to self | + | *‘Number one. [pause] The most important reason. That’s going to be the risk to myself.’* P02 |
|  |  | Perceived increased risk due to environmental change | + | *‘I think just with everything changing, the weather fluctuating and climate change, I think for me at least, I tend to catch a cold easily with the weather change. So I don’t want to take risks with any of the respiratory related health issues.‘* P12 |
|  |  | Perceived severity of TB risk in high-risk settings | +/- | ‘You could be looking after someone with TB, and you're already a dormant TB, that’s a positive, so that put me probably a little more at risk that someone who doesn’t have, yeah.’ P04  *‘I think there is a role for preventative antibiotics, especially for individuals who probably can identify the possibility of exposure, especially coming from countries or going to countries that have a high incidence of TB. So having a lower risk of contracting TB versus having a high risk of contracting TB.’* P16 |
|  |  | Perceived severity/lack of severity of future TB risk | + | *‘I mean, the risk seems very low. It's one in 100.’* P07  *‘Well, if the average is 2%, so anything above 5% then that’s really scary, isn’t it?’.* P05 |
|  |  | Uncertainty of when TB will become active | + | *‘The reason why it worries me to a point is the fact that, I say something that I really do not like to give a false sense- Uncertainty. Anything can happen at any time. Yeah, anything can happen at any time and you don’t want to pose a risk to yourself and pose a risk to others, so that’s basically it honestly.’* P02 |
| 8. Intentions (A conscious decision to perform a behaviour or a resolve to act in a certain way) | 1. Commitment to take treatment  Participants expressing a firm intention to take preventive treatment, including maintaining commitment despite uncertainty about treatment effectiveness or changes in future TB risk. | Commitment to take treatment despite uncertainty of effectiveness | + | *‘I do (think taking the antibiotics will help stay healthy). And even if it doesn't, at least I can say I tried.’* P06 |
|  |  | Commitment to take treatment | + | *‘If that medicine can control it then I’ll take it. […] If I need to take them for me to be healthy, like that’s easy. I’m going to do it’.* P03  *‘I’m 100% ready for it. I’m happy to take it.’* P12 |
|  |  | Maintain intention to take treatment despite variation in future risk | + | *‘I’m still going to take the preventative antibiotics.’* P02  *‘Yes, as long as it will prevent the disease. [question: even if the risk was very low?] Yeah- […] To me it’s low but you don’t know some day.’* P03  *‘So, yes, if it is going higher, definitely will push me towards taking the medication, but if it is actually going lower, I don’t think it will have any impact on my decision to take the medication or not.’* P05 |
|  | 2. Conditional intention to take treatment  Participants expressing willingness to take preventive treatment under specific conditions, such as attendance to follow-up testing or if the TB risk to themselves or others increased. | Willingness to take treatment due to follow up testing | + | *‘The only way I'm going to be okay with it is because every two weeks, I've got to do what they say, they'll be checking the blood test.’* P17 |
|  |  | Willingness to take treatment if risk for self and others increases | - | *‘Yeah, if I had a higher risk, then I would definitely take treatment.’* P10  ‘*I have to think of where I’m living, if it’s going to be a risk to people around me and others then I will take it.’* P20 |
|  | 3. Intention to decline or stop treatment if it becomes a burden  Participants' intentions not to start, or to stop, preventive treatment if perceived to be too burdensome, particularly when it disrupted daily activities or routines. | Intention not to take the treatment | - | *‘My intention is actually not taking it up to now.’* P07 |
|  |  | Intention to stop treatment if they disrupt daily activities | - | *‘If I’m experiencing too much fatigue [unclear 00:17:23] and if it’s affecting my daily activities, I think I’d stop the medication.’* P13 |
| 9. Goals (Mental representations of outcomes or end states that an individual wants to achieve) | 1. Long term health is a priority  Goal of promoting their own health, including protecting long-term health and preventing progression to active TB. | Long-term health focus | + | *‘If I have no other medical condition, I would have not but that’s basically- So what, you know, I must take medication because I, got other medication for- […] It’s something to do with my heart condition. [ …] I don’t want my heart to be burdened by TB in the future’.* P04  *‘I think it is not the risk of TB actually that made me to take the medication, it’s the risk of the possible things that could happen to me in the future. […] If there is any chance it’s going to help your future health, if that makes sense, then why not do it?’* P05 |
|  |  | To prevent active TB | + | *‘Yeah, that’s it, for my family, for me to be healthy, for me not to have that disease because I don’t want that disease’.* P03  *‘It's all about to not get TB in the future.’* P21 |
|  | 2. Priority to prevent transmission  Goal of preventing TB transmission to others, including protecting family and the wider society. | | + | *‘To prevent the disease, not having, like to spread it, because if you have it, you’re going to spread it’.* P03  *‘I think the most important reason would be family.[…] I feel like if I can prevent them from getting it, by any means possible, I will. I will do whatever it takes.’* P06 |
| 10. Memory, attention and decision processes (The ability to retain information, focus selectively on aspects of the environment and choose between two or more alternatives) | 1. Difficulty to remember to take treatment  Participants anticipating challenges in remembering to take preventive treatment consistently. | | - | *‘thinking that I have to wake up and take it every day, not forgetting.’* P06  *‘Sometimes you might just forget and this and that.’* P25  *‘I’m pretty sure I would not remember to take the medication.’* P16 |
|  | 2. Knowing you can change your mind  Participants taking into account that the decision to accept preventive treatment is not fixed and could be revised later if their views or circumstances change. | | - | *‘I decided not to have it now was they gave me the option, like if I thought in the future or if I think that I need it in the future, I can always come in. […] I’m lucky I have the option to change my mind.’* P01 |
|  | 3. Time to discuss process and take decision  Participants taking time to discuss the treatment and consider their options before making a decision. | | - | *‘instead of asking for people to make an in-prompt decision […] Because in two minutes we have together, […] It’s not sufficient for me. But I think the better way would have been […] Here’s the information that you can look until come back to us with a decision whether you want to take treatment […] Yeah, no, but so I have to be honest, it’s helped me as well, this questioning or whatever was spoken now. It gave me a second thoughts as well, honestly, because some of the things that we’ve just dealt with they would never have crossed my mind, not until we spoke of them now.’* P01  *‘I need to take my time to think, think it through. So I’m still in the thinking stage.’* P19 |
| 11. Environmental context and resources | 1. Accessibility of care for active TB reduces engagement for preventive treatment | Having quick access to treatment for active TB if symptoms start | - | *‘whenever I feel this kind of symptom immediately I can call the emergency number. That’s a very good thing that if I’m affected with this kind of infection, I can get immediate treatment which is very good for me.’* P10 |
|  |  | Monitoring TB progression | - | *‘Once I come back from my other consultation, I’ll see if the virus level is lowering or getting bigger. So to be honest nothing right now scares me.’* P08 |
|  | 2. Busy environments cause non-adherence  Participants described how busy or demanding environments (e.g., urban settings, eating patterns, lifestyle demands, and work constraints) make it harder to consistently adhere to treatment. | Urban environments | - | *‘it also affects my time schedule as in London people are very hurried for their schedules so yeah.’* P10 |
|  |  | Eating patterns | - | *‘I don’t think it will impact anyway unless if it’s an antibiotic that I have to eat before using because normally I do intermittent fasting.’* P25  *‘because I don’t normally eat three times. So besides that, if it is one a day, I don’t mind, it will be fine.’* P12 |
|  |  | Lifestyle constraints | - | *‘If I’m moving around to different places. I’m probably going to forget.*  *Yeah, if I’m away from home. Honestly, just if I’m busy.’* P14 |
|  |  | Work constraints | - | ‘Because I’m on nights tomorrow up until Sunday, and I don’t want to forget that- You know, to take the medication.’ P04 |
|  | 3. Environmental risk context impacts engagement for preventive treatment  Participants discussed environmental context, including environmental changes and factors that increased perceived risk of exposure or reinfection. | Environmental changes and new health concerns | + | *‘I think just with everything changing, the weather fluctuating and climate change. […] because we’re all falling sick quite often lately because of the weather change’* P13 |
|  |  | Environmental factors that increase risk of reinfection | +/- | *‘I’m thinking if I finish my training, I might want to go back home and it’s quite an endemic zone there, so I might get re-infect again, so what is the point of taking the antibiotics?’* P05  *‘Because I’m still working in [workplace] and within an infectious- So I mean, who knows, it might be dormant now, you know, I’m always exposed to this.’* P04  *‘The most important thing to consider taking the treatment would be if you are actually exposed to high risk population, that’s what I would say.*’ P11 |
|  | 4. Mixed views on the need to access hospitals to get and monitor treatment  Participants expressed mixed views about hospital-based care for preventive treatment, with follow-up visits and tests seen by some as reassuring, while others discussed a lack of timely or local medical support and viewed multiple hospital visits as a barrier to engagement. | Follow up and tests as reassuring | + | *‘I’m not so bothered and the fact that I’m having a blood test now and in the next two weeks when I start, when I start the antibiotics and two weeks after starting the antibiotics […] So I’m not so bothered, you know?’* P02  *‘there’s a follow-up, they will call you, on how you’re feeling. So there’s a follow-up so they will tell you if you have maybe come and do a test. So I think the two weeks check-up, those are the reasons. So if you feel you are not okay and all this, when they check you up they will know what is happening. So I feel that is why there is a two week checkup.’ P09* |
|  |  | Lack of timely and local medical support | - | *‘if I notice a side effect and not being able to get a GP appointment quick enough or seeing somebody quick enough or even getting blood tests if I need to get blood tests done quick enough.’* P25  *‘In terms of the frequent blood tests, it would have to be a case where I can get the blood tests done in another trust or through my GP practice in an area that is closest to me so that saves them coming directly to the hospital to get blood tests done so that at least I can probably get the tests done during lunchtime or right before work if possible which would be easier for me.’* P16 |
|  |  | Multiple hospital visits as a barrier | - | *‘I mean, if I had to like basically come back to the hospital again and again to get the medications.’* P08  *‘I might be starting sometime next week because I have to be here again in the next two weeks of which I don’t think I’m going to be in London.’* P02  *‘I’m here and apparently there’s no medication so I’ll have to come back again. […] and it’s a whole other medical treatment process and it’s exhausting to access treatment, travel and all of that.’* P13 |
|  | 5. Treatment characteristics as barriers to adherence to treatment  Participants described features of the treatment as barriers to engagement, including dose frequency, medication properties, side effects, and treatment duration. | Dose frequency | - | *‘if it is like maybe a one-time medication, just take it once and that’s it, that would be actually much easier but having to deal with it for three months. […] Well maybe the medication can maybe like a weekly dose that might also help’.* P05  *‘or having a medication that you don’t have to take every day or even an injectable option that you take on a weekly basis, I don’t know.’* P16  *‘I think I would consider it as more likely because maybe in taking it I don’t have to be regular. Maybe the injections last for a couple of months.‘* P20 |
|  |  | Medication properties | - | *‘The size of the tablets I guess if they’re difficult to swallow, it would make it quite difficult.’* P14  ‘- the taste, the size I think.’ P19  *‘I could consider taking an injection and see how that would affect me or some other forms of treatment but taking drugs, I don’t know.’* P20 |
|  |  | Type of side effects | - | *‘I mean I’m not too sure of the current medication, if it causes GI side effects but having a medication that doesn’t really affect the GI system. […] It would have to be a medication that doesn’t affect the GI system that much.’* P25 |
|  |  | Treatment duration | - | *‘ So it would have to be a shorter duration, not three months. Well one month or six weeks would be fine.’* P25 |
| 12. Social influences (Those interpersonal processes that can cause individuals to change their thoughts, feelings, or behaviours) | 1. Practical and emotional support from family and friends to the decision to take and subsequent adherence  Participants described practical and emotional support from family and friends as helping both with deciding to start preventive treatment and with taking treatment as prescribed. | | + | *‘I told my wife about it but she was in support of it also. […] She felt taking the medication would be the best so she was in support of it also. We came to the conclusion that I should just go for it.‘* P09  *‘I think one thing that might help me, that’s going to make it easier, you know, it’s a bit of support in terms of support from family. Like I could, actually, my wife could actually support and encourage.’* P02 |
|  | 2. Social pressure and norms that influence initiation  Participants described how social pressures and norms shaped decisions about starting preventive treatment, including anticipated stigma, influence from others’ experiences of TB and its treatment, as well as interactions with vulnerable people. | Anticipated stigma | - | *‘So you don’t even know how to explain to people that are close to you like, yeah, they said that I’m not infected but I’m still taking the medication’.* P01  *‘and then if you want to travel somewhere, it's also some stigma associated with taking medications. So those all have to be counted in.’* P11 |
|  |  | Cultural/social norms | - | ‘Some people would have religion, like religious belief or cultural belief or- More than socioeconomic background could actually be the reason. Some people back home, there are some people that believe that they shouldn’t take drugs because drugs are kind of harmful’. P02 |
|  |  | Information from others about their experience | + | *‘I found some people with same latent, you know, TB that haven’t really developed into the active stage’.* P02  *‘I've seen first hand experience. My girlfriend is taking the same medication and it has actually made her much better.’* P08  *‘I know of cases where friends or relatives have had some form of medication for treatment and because of side effects they’ve had to take others and then it led to many other complications.’* P20 |
|  |  | Living or working with vulnerable people | + | *‘If I were working with children, if I had my own children at any point or if I were living with elderly population who are at greater risk than getting an infection […] I might consider.’* P11 |
|  | 3. Trust in healthcare and medicines  Participants’ trust in healthcare professionals, prescribed treatment, and the wider medical system shaped engagement with preventive treatment. | Lack of trust in pharmaceutical industry | - | *‘You know that there’s a bit of a scare when it comes to modern medicine. You know, there are different theories about big pharma companies trying to do this, trying to do that.’* P19 |
|  |  | Health professionals’ "sales pitch" can impact treatment decisions | - | *‘And also, whoever is prescribing this medicine, they are the expert in this field. So they definitely know better than me. So they are giving the choice, and I will definitely trust them if I have to.’* P08 |
|  |  | Support from health professionals for active TB | - | *‘at the same time I know the NHS staff are always there to support me so there is no worry.’* P10 |
|  |  | Support from health professionals to understand latent TB and its treatment | + | *‘I think the information was quite adequate and I was told that my chest x-ray, everything was normal. I don’t have any active disease at the moment, so I can take the medication and I can decide not to take it, but I was given the reasons why I should take it and also if I prefer not to take it this might be the consequences. So basically, I think the information I got from the hospital was quite adequate.’* P01 |
|  |  | Trust in the healthcare professionals and prescribed treatment | + | *‘I don’t want to think that it won’t work because they wouldn’t give it to me unless it’s proven. [ …] Yeah, why do you do consultation if you don’t listen to the doctor?’* P03  *‘And also, whoever is prescribing this medicine, they are the expert in this field. So they definitely know better than me. So they are giving the choice, and I will definitely trust them if I have to.’* P08 |
| 13. Emotion (A complex reaction pattern, involving experiential, behavioural, and physiological elements, by which the individual attempts to deal with a personally significant matter or event) | 1. Fear of the consequences of not taking the treatment  Participants described fear and worry about the potential consequences of not taking preventive treatment, including concern about developing active TB in the future and missing symptoms, and living in environments perceived to increase risk. | Fear of getting active TB in the future | + | *‘Because again it’s something that scares me a bit. Because if I get sick, there’s other factors which might change my life, my other stuff and my hobbies.’* P09 |
|  |  | Fear of missing active TB symptoms | + | *‘So now I’m not only thinking, like I’m feeling better in myself, what if- The fact is I’m not really well, but I was, I couldn’t identify that or I couldn’t tell myself and I jeopardise my family from not taking the treatment.’* P05 |
|  |  | Fear of what could happen in the future | + | *‘Yeah, like I said, it was just the fear of what’s going to happen in the future, nobody can see the future so you just have to start planning now.’* P05 |
|  |  | Worry of being in a high risk environment | + | *‘Because me, I travel home like twice a year and I stay like a couple of weeks to a month, you know? And knowing what I’m doing, I’m very much among people. I’m just like very secluded, no, I always stay among at least some people. So having this at the back of my mind is getting me worried now, honestly.’* P01 |
|  | 2. Fear of the consequences of taking the treatment  Participants described fear and anxiety about the potential consequences of taking preventive treatment, including concerns about minor or serious side effects and the duration of treatment, worries about taking too much medication, and possible effects on underlying conditions. | Fear/anxiety about uncertainty (making the right decision taking the treatment) | +/- | *‘I mean even though I have a nursing background, there’s always anxiety, you know? Am I making- You know, even now, I’m thinking, am I making the right decision* [taking the treatment] *even though I’m [unclear 00:22:38] this, but, you know? There’s always like a query’* P04 |
|  |  | Fear of minor side effects | - | *‘I think I’m not going to be worried about the major side effects but just the minor ones. I think my body might experience those. […] I think my body usually feels tired or experiences fatigue when taking antibiotics and that definitely affects my energy levels and how I go about the day, I’m a little worried about that.’* P13 |
|  |  | Fear of serious unpredictable or unusual side effects | - | *‘Especially if other side effects come up as well, I’d probably get really scared and probably think that it’s irreversible and not wanting to have any adverse complications from the medication.’* P16 |
|  |  | Fear of taking too much medication | - | *‘Maybe they’re scared.[…] Scared of taking too much medicine’.* P03 |
|  |  | Fear of treatment effects on underlying condition | +/- | *‘But some people actually fear that they could be more complicated and the health status of some people might actually- Might actually discourage that because- […] I believe that some people’s health status, I’d say probably those with the underlying sickness at the moment […] Probably if they take the antibiotics, that could actually, you know, still affect them if they have an underlying condition?’.* P02 |
|  | 3. Low mood associated with diagnosis and treatment  Participants described low mood and emotional distress related to TB diagnosis and preventive treatment. | Feeling upset by lack of reactivity from work when exposed to TB | +/- | *‘Yeah, I've got it through them. Because I had contact with someone who has a TB, and I was really angry, because I think maybe they didn't figure it out on time. Like the person is going through TB. So the thing is, I'm upset because the kind of job I do, I'm trying to care for people. It's like you're taking risks for your health wise, because that's not a nice thing.’* P17 |
|  |  | Low mood following TB test results | +/- | *‘Okay, initially like I said I was a bit down.’* P02  ‘There’s an initial worry, definitely.’ P04 |
|  |  | Low mood having to take a treatment | - | *‘Because I think, to me it’s got like a bit of, er, I don’t know, like within my thinking I’m feeling like when once I take it, or if I decided taking it, I will feel very, very down.’* P01 |
|  | 4. Reassurance associated with clear information and planning  Participants described feeling reassured when latent TB and its treatment were clearly explained and when there was a defined plan for treatment. | Feeling reassured by treatment plan | + | *‘If I haven’t got a treatment plan, that would have weighed me down. So there’s a treatment plan for that for me so it’s fine with me.’* P12 |
|  |  | Feeling reassured when latent TB explained | + | *‘sometimes you may not feel good about your test but when you get a proper explanation about everything that is going on, you tend to calm down. At first I was not happy but when I got a proper explanation and proper, he educated me on everything so I said okay, that’s good.’* P09 |
| 14. Behavioural regulation (Anything aimed at managing or changing objectively observed or measured actions) | 1. Strategies to monitor and manage side effects  Participants described strategies to monitor and manage side effects and plan in advance to attend check-ups. | Monitoring and planning in advance for managing and responding to side effects | + | *‘So that’s why Monday when I’m off, I’ll start taking that medication rather than, you know, starting today or tomorrow when I’m actually working and, you know? But if there’s some side effects so-’* P04 |
|  |  | Plan in advance for check-ups | + | *‘it will do with my schedule of work. So if I have the information ahead about the time, I don’t mind coming.’* P12 |
|  | 2. Strategies to initiate and adhere to treatment  Participants described strategies to start and maintain preventive treatment, including fitting treatment around existing routines or creating new ones, and timing treatment initiation to accommodate practical constraints. | Adapt treatment to eating patterns | + | *‘Well, again yes, because I was just wondering because I do fast some few days in a week. So now I’m just wondering, okay, what will happen on the days that I’m fasting, will I have to take them early in the morning or when I break my fast […] But I’m sure I’ll find a way around it.’* P05 |
|  |  | Adhere to treatment using existing medication routines | + | *‘Because I’m taking medication already so I know when to sort of like- I know my way how to remind myself to take medication.’* P04 |
|  |  | Create routine to adhere to treatment | + | *‘I wouldn't really say the time. I'd say more of the routine, like having that routine of taking the treatment every day, thinking that I have to wake up and take it every day.’* P06  *‘So if you are doing something every day, I don’t think you’ll have any issue with it. This is what I do every day. You should be familiar with it so you shouldn’t have any issues with it.’* P09 |
|  |  | Identifying strategies to help take medication as required | + | *‘you know, set off alarms and all of that. You haven’t taken your drug today.’* P02  *‘I would probably just put them next to, I don’t know, my toothbrush and just take it every morning when I brush my teeth or every evening or whatever.’* P14 |
|  |  | Timing treatment start to fit with practical constraints | + | *‘I might be starting sometime next week because I have to be here again in the next two weeks […] So, I’m going to start taking it to fit into the time when I’m going to be in London, like in two weeks.’* P02  *‘I’m trying to take it, it starts Monday, just to give timing. Because I’m on nights tomorrow up until Sunday, and I don’t want to forget that- You know, to take the medication.’* P04 |
|  | 3. Alternative strategies to manage the risk of active TB without treatment  Participants described alternative approaches to managing TB without preventive treatment, including changing habits to reduce risk and self-monitoring for symptoms of active TB. | Change habits to limit risk of active TB | - | *‘No, not now because as I told you, I changed so many dietary, I do my dietary changes and turning my life towards a healthy life so there is no worry, yeah.’* P10 |
|  |  | Self-monitoring for symptoms of active TB | - | *‘So that’s why I think, well, I can certainly wait and see this time. [ …] Now I know the symptoms that I should be looking for, if I start feeling those kind of way, I think I have them, I start feeling like loss of weight or intractable cough, you know what, I think I need to come for a blood test or I need get some imaging.’* P01  *‘I would think it would be the best to wait and see if I do develop any symptoms or if I do have any active infection or something that I can look for.’* P11 |
